# Dietary Intake Is Associated With the Prevalence of Uterine Leiomyoma in Korean Women: A Retrospective Cohort Study

**DOI:** 10.1101/2023.08.23.23294524

**Authors:** Min-Jeong Kim, Sunmie Kim, Jin Ju Kim, Young Sun Kim, Ji Hyun Song, Jung Eun Lee, Jiyoung Youn, Sun Young Yang

**Author notes:** These corresponding authors contributed equally to this work Sunmie Kim, M.D. Ph.D. Department of Obstetrics and Gynecology, Seoul National University College of Medicine, Seoul National University Hospital Healthcare System Gangnam Center, 39FL., Gangnam Finance Center 152, Taeheranro, Gangnam-gu, Seoul, 06236, South Korea; Sun Young Yang, M.D. Ph.D. Department of Internal Medicine, Division of Gastroenterology, Healthcare Research Institute, Seoul National University Hospital Healthcare System Gangnam Center, 39FL., Gangnam Finance Center 152, Taeheranro, Gangnam-gu, Seoul, 06236, South Korea.

## Abstract

**Objective:** Uterine leiomyoma (UL), the most prevalent benign gynecologic tumor among reproductive-aged women, lacks sufficient research on the potential association between dietary intake and its occurrence in Korean women. Addressing this research gap, this study aims to evaluate the potential link between dietary intake and the prevalence of UL in Korean women.

**Methods:** In this cross-sectional study, a cohort of 672 women, aged 25 to 65, were enrolled, with 383 (57%) being premenopausal. Dietary intake was assessed using a validated food frequency questionnaire (FFQ), and UL presence was determined through ultrasonography. The analysis focused exclusively on items within ten categories, including vegetables/fruit, vegetables, fruits, red meat, processed meat, poultry, fish, dairy product, milk, and alcohol. Multiple logistic regression models were employed to explore the relationship between dietary intake and the prevalence of UL, calculating odds ratios (ORs) and 95 % confidence intervals (CIs) while adjusting for confounding factors.

**Results:** Within the total cohort, 219 (32.6%) women were diagnosed with UL. High intakes of fish and poultry showed an association with higher UL prevalence. Odds ratios (95% CIs) for the upper quartiles compared to the lower quartiles were 1.70 (1.02-2.84; p trend = 0.049) for fish intake and 1.85 (1.09 -3.14; p trend = 0.07) for poultry intake. Conversely, an inverse relationship emerged between dairy product intake and UL prevalence, with an odds ratio of 0.59 (95% CI 0.36–0.98; p trend = 0.06). Stratifying the analysis by menopausal status revealed a parallel pattern, with heightened UL prevalence with fish intake and reduced prevalence with dairy product intake. However, the link between poultry intake and UL prevalence was primarily observed among postmenopausal women. Among premenopausal women, elevated vegetable intake was linked to a decreased UL prevalence (OR 0.47, 95% CI 0.22-1.01 for top vs. bottom quartiles; p trend = 0.01).

**Conclusion:** We found that high consumption of fish and poultry, coupled with low intake of dairy products, correlated with an elevated prevalence of UL. Furthermore, vegetable intake exhibited an inverse relationship with UL prevalence, particularly among premenopausal women.

## Introduction

Uterine leiomyoma (UL) is the most common benign gynecologic tumor, affecting approximately 25% of women of reproductive age, with peak prevalence occurring at age 50 and a lifetime risk of up to 70% [1-3]. While the pathophysiological mechanisms underlying the development of ULs at the cellular and molecular level have not been fully elucidated, they appear to be sex-hormone (estrogen and progesterone) dependent diseases, typically appearing after menarche, growing during reproductive ages, and regressing along with declining reproductive hormone levels after menopause [4-7]. Other known associated factors include age, ethnicity (with 2-3 times higher incidence in black women than in other races), genetics, number of pregnancies (more common in women who have had fewer pregnancies or deliveries), obesity, lack of physical exercise, and some dietary factors [8, 9].

As data on the relationship between dietary factors and malignant diseases such as breast or endometrial cancer, which are presumed to be estrogen dependent, have been reported mostly in terms of the potential of chemoprevention and long-term prognosis [10-13], the role of dietary nutrition as a factor that can be modified in the development and growth of UL has become a topic of interest, as dietary intake may alter either endocrine function or molecular biologic milieu [14].

According to previous studies, dietary patterns or some nutrients have shown significant associations with ULs. While the consumption of fruits and vegetables has shown a protective effect against ULs, findings have been inconsistent for other foods such as dairy, meat, or fish [8, 15-17]. Meanwhile, studies that have reported the association between nutritional intake analysis and the prevalence of UL in Korean women are limited. This study aimed to investigate the association between dietary intake and prevalence of UL stratified by menopausal status among Korean women who underwent both analysis of food intake and pelvic ultrasound exam from a previous cross-sectional study of our institute.

## Materials and Methods

### Study design and participants

This study retrospectively used a prospectively collected cohort from our previous study [18]. Participants who underwent health checkups, including colonoscopy and dietary intake assessment, using a semi-quantitative food-frequency questionnaire (FFQ) at the Seoul National University Hospital Gangnam Center in Seoul, Korea, between May and December 2011, were registered. Among them, only women participants who also had pelvic ultrasonography during the study period were enrolled. Individuals who had already undergone hysterectomy or who did not take the pelvic ultrasound examination were excluded. A total of 672 women were finally included in this study. This study was approved and has been granted an exemption from the requirement for additional consent procedures by the Institutional Review Board (IRB) of this institution. This exemption is based on the fact that the study involves the analysis of medical records from women who underwent pelvic ultrasound examinations among the participants of a previously conducted study at our institute [18]. To ensure the protection of personal information of the subjects included in this study, participants’ names were anonymized, and participant identification codes and medical record numbers were encrypted. Access to the data of the study participants was conducted for one year, from June 2015 to May 2016.

Postmenopausal status was defined as the absence of menstruation for at least 1 year. Women in peri-menopausal status (irregular cycles of more than ≥7 days differences or missed two or more cycles of menstruation within 12 months) were classified as premenopausal women [19].

### Clinical and laboratory assessment

Baseline characteristics, such as medication use (e.g., antidiabetic, antihypertensive, or lipid-lowering agents), underlying diseases (diabetes, hypertension, and dyslipidemia), smoking history, amount of physical activity, alcohol consumption, and reproductive characteristics (age at menarche, parity, age at first delivery, and menopausal status) were recorded during a medical interview using a structured questionnaire before a routine gynecologic examination. Anthropometric parameters (body mass index (BMI), waist circumference (WC), and blood pressure (BP)), and biochemical results (fasting plasma glucose, triglycerides, low-density lipoprotein (LDL)-cholesterol, and high-density lipoprotein (HDL)-cholesterol) were retrospectively reviewed for each individual, as previously described.

### Assessment of uterine leiomyoma

ULs were assessed through ultrasound examination using GE LOGIQ^®^9 (GE healthcare, General Electric Co., UK) equipment. The examination was performed by one of the three gynecologists who were obstetrics and gynecology specialists (M-J Kim, JJ Kim, and S Kim) with more than eight years of experience. The presence of UL was assessed only by intracavitary (mostly transvaginal, some transrectal) pelvic ultrasound examination, and cases with UL were defined as having one or more nodules of typical leiomyoma with ≥10mm in length.

### Assessment of Dietary intake

Dietary intake data were assessed prior to the examination on the same day using a validated 106-item Food Frequency Questionnaire (FFQ) [20] with assistance from a registered dietician. Participants reported their usual frequency of consumption of various foods and typical portion sizes for the year preceding the interview date. Each food item had 9 options for frequency (ranging from “never or less than once per month” to “3 times per day”) and three options for portion size (‘small”, “medium”, or “large”). Fruit and vegetable intake included all raw, cooked, canned, frozen or dried forms of fruits and most edible vegetables. For the analysis, we examined the food consumption and total energy intake. Only items corresponding to the ten categories (vegetables/fruit, vegetables, fruits, red meat, processed meat (grouped into tertiles, two categories in postmenopausal women), poultry, fish, dairy product, milk, and alcohol (grouped into tertiles)) were included in the analyses and the amount of food intake was divided into quartiles.

### Assessment of risk factors

Metabolic syndrome (MetS) was defined according to the harmonized definition proposed by the International Diabetes Federation/American Heart Association/National Heart, Lung, and Blood Institute [21]. A patient was diagnosed with MetS if they met three or more of the following criteria: abdominal obesity (waist circumference ≥85 cm for Korean women as proposed by the Korean Society for the Study of Obesity [22]), high triglycerides (TG) (≥ 150 mg/dL), low HDL-cholesterol (< 50 mg/dL), high fasting glucose(≥100 mg/dL) or treatment for diabetes, and increased blood pressure (≥ 130/85 mmHg) or treatment for hypertension.

Current smokers were defined as those who had been smoking at least one cigarette per day during the previous 12 months, while past-smokers were considered those who discontinued smoking for at least 12 months before inclusion in the study. “Ever smokers” refers to respondents who are current or past smokers.

Physical activity (PA) was measured by the modified Korean version of the PA questionnaire from the National Health and Nutrition Examination Survey [23]. PA was quantified using metabolic equivalent (MET)-minutes per week.

### Statistical analysis

Numerical variables were expressed as mean±standard deviation, and categorical variables were presented as numbers and percentages. If the parameters were not normally distributed, log_10_ transformation was used for analysis. The relationship between each dietary intake and UL was evaluated using binary logistic regression analyses. As ULs usually shrink after menopause due to a drastic drop in serum estrogen levels, the data were analyzed separately for two groups based on their menopausal status (premenopausal, including peri-menopausal, vs. postmenopausal). The median value of each tertile or quartile was included in the models as a continuous variable for trend testing. Odds ratios (ORs) and 95% confidence intervals (CIs) were calculated to evaluate the associations using multiple logistic regression models. In Model 1, we adjusted for confounding variables including age (years, continuous), BMI (kg/m^2^, <18.5, 18.5–23, 23–25, 25≤), total energy intake (kcal/d, quintile), and LDL-cholesterol (mg/dL, continuous). In Model 2, we further adjusted for all clinically relevant parameters, including menopausal status (premenopausal vs. postmenopausal), age at menarche (years old, ≤11, 11<), age at first delivery combined with parity (nulliparity, years old, <25, 25 ≤), alcohol intake (g/d, continuous), smoking status (never, or ever) and physical activity (MET-min/week, tertile). All analyses were performed using SAS 9.3 (SAS Institute Inc., Cary, NC, USA). We used 2-sided statistical tests, and *p*-values less than 0.05 were considered statistically significant.

## Results

### Baseline characteristics and prevalence of UL

A total of 672 women were included in the study, with 383 (57%) being premenopausal and the age range being 25–65 years old (mean age 50.1 years). Of these, 219 (32.6%) were diagnosed with UL, with no significant difference in prevalence between pre-and postmenopausal women (34.5% vs. 30.1%, respectively; *p* = 0.23). Compared to women without UL, those with UL were older (51.0 ± 7.4 vs. 49.7 ± 9.5 years, *p* = 0.01), had a higher BMI (22.4 ± 3.2 vs. 21.9 ± 2.8 kg/ m^2^, *p* = 0.03), and higher LDL-cholesterol levels (128.2 ± 31.3 vs. 122.3 ± 32.4 mg/dL, *p* = 0.01). There were no significant differences in terms of reproductive, lifestyle, comorbidities, or laboratory parameters between the two groups (Table 1).

**Table 1.**
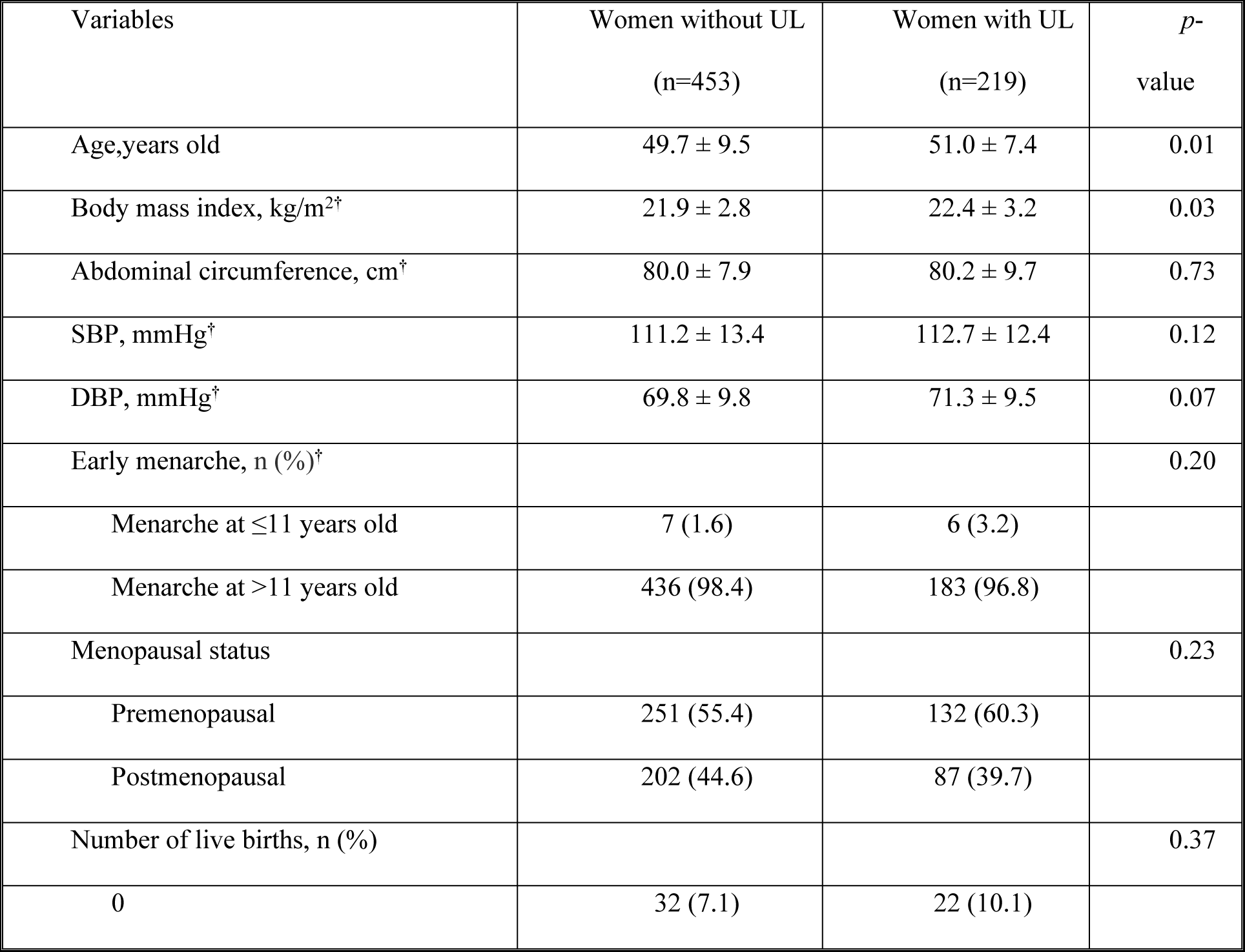
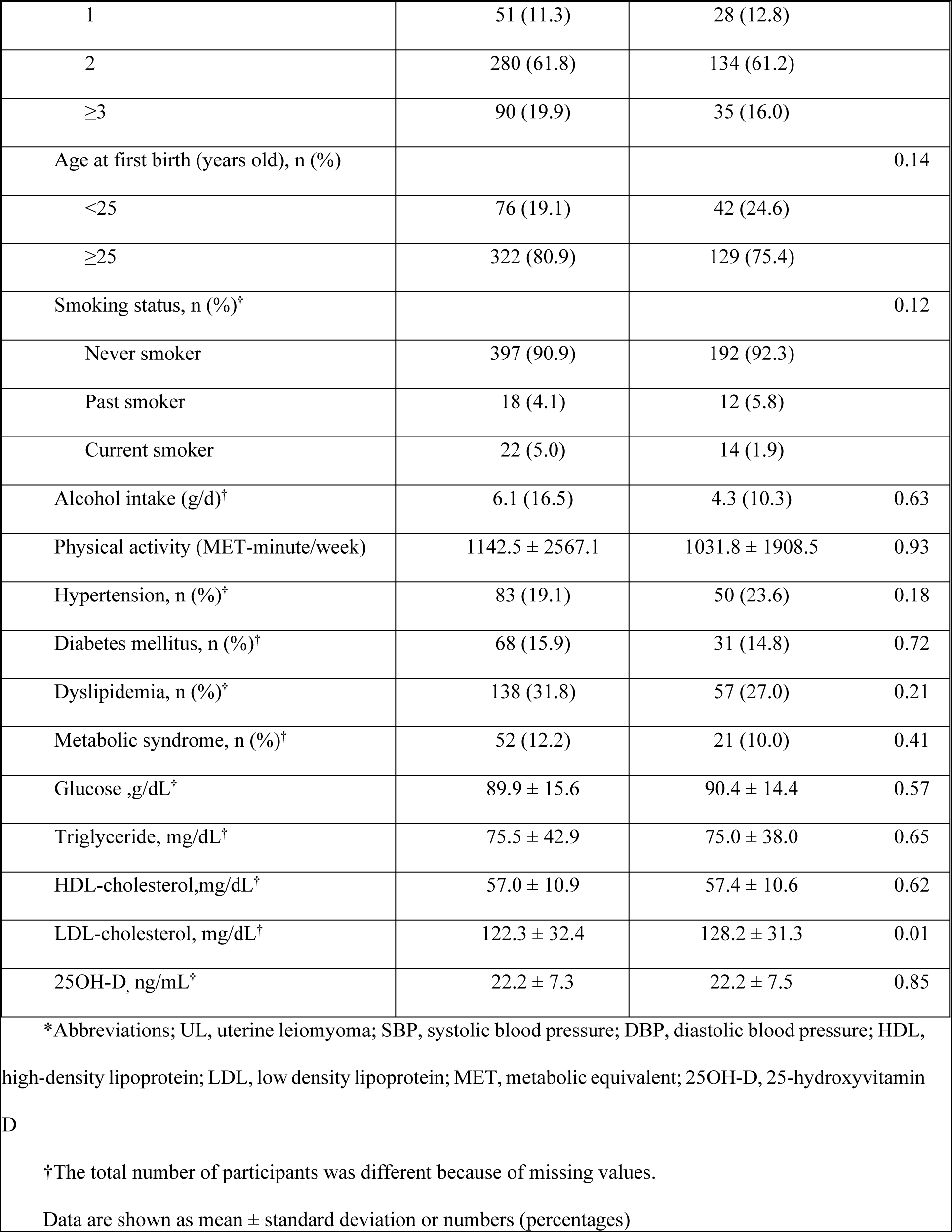
Baseline characteristics of participants with and without uterine leiomyoma.

| Variables | Women without UL<br>(n=453) | Women with UL<br>(n=219) | *p*-<br>value |
| --- | --- | --- | --- |
| Age, years old | 49.7 ± 9.5 | 51.0 ± 7.4 | 0.01 |
| Body mass index, kg/m<sup>2</sup>† | 21.9 ± 2.8 | 22.4 ± 3.2 | 0.03 |
| Abdominal circumference, cm† | 80.0 ± 7.9 | 80.2 ± 9.7 | 0.73 |
| SBP, mmHg† | 111.2 ± 13.4 | 112.7 ± 12.4 | 0.12 |
| DBP, mmHg† | 69.8 ± 9.8 | 71.3 ± 9.5 | 0.07 |
| Early menarche, n (%)† |  |  | 0.20 |
| Menarche at ≤11 years old | 7 (1.6) | 6 (3.2) |  |
| Menarche at >11 years old | 436 (98.4) | 183 (96.8) |  |
| Menopausal status |  |  | 0.23 |
| Premenopausal | 251 (55.4) | 132 (60.3) |  |
| Postmenopausal | 202 (44.6) | 87 (39.7) |  |
| Number of live births, n (%) |  |  | 0.37 |
| 0 | 32 (7.1) | 22 (10.1) |  |
| 1 | 51 (11.3) | 28 (12.8) |  |
| 2 | 280 (61.8) | 134 (61.2) |  |
| ≥3 | 90 (19.9) | 35 (16.0) |  |
| Age at first birth (years old), n (%) |  |  | 0.14 |
| <25 | 76 (19.1) | 42 (24.6) |  |
| ≥25 | 322 (80.9) | 129 (75.4) |  |
| Smoking status, n (%)<sup>†</sup> |  |  | 0.12 |
| Never smoker | 397 (90.9) | 192 (92.3) |  |
| Past smoker | 18 (4.1) | 12 (5.8) |  |
| Current smoker | 22 (5.0) | 14 (1.9) |  |
| Alcohol intake (g/d)<sup>†</sup> | 6.1 (16.5) | 4.3 (10.3) | 0.63 |
| Physical activity (MET-minute/week) | 1142.5 ± 2567.1 | 1031.8 ± 1908.5 | 0.93 |
| Hypertension, n (%)<sup>†</sup> | 83 (19.1) | 50 (23.6) | 0.18 |
| Diabetes mellitus, n (%)<sup>†</sup> | 68 (15.9) | 31 (14.8) | 0.72 |
| Dyslipidemia, n (%)<sup>†</sup> | 138 (31.8) | 57 (27.0) | 0.21 |
| Metabolic syndrome, n (%)<sup>†</sup> | 52 (12.2) | 21 (10.0) | 0.41 |
| Glucose ,g/dL<sup>†</sup> | 89.9 ± 15.6 | 90.4 ± 14.4 | 0.57 |
| Triglyceride, mg/dL<sup>†</sup> | 75.5 ± 42.9 | 75.0 ± 38.0 | 0.65 |
| HDL-cholesterol,mg/dL<sup>†</sup> | 57.0 ± 10.9 | 57.4 ± 10.6 | 0.62 |
| LDL-cholesterol, mg/dL<sup>†</sup> | 122.3 ± 32.4 | 128.2 ± 31.3 | 0.01 |
| 25OH-D, ng/mL<sup>†</sup> | 22.2 ± 7.3 | 22.2 ± 7.5 | 0.85 |

\*Abbreviations; UL, uterine leiomyoma; SBP, systolic blood pressure; DBP, diastolic blood pressure; HDL, high-density lipoprotein; LDL, low density lipoprotein; MET, metabolic equivalent; 25OH-D, 25-hydroxyvitamin D
<sup>†</sup>The total number of participants was different because of missing values.
Data are shown as mean ± standard deviation or numbers (percentages)

### Association between dietary intake and prevalence of UL in the entire population

Table 2 shows the associations between dietary intake and the prevalence of UL in all participants, as analyzed through age-adjusted and two-stage multiple logistic regression models. Among all participants, higher fish intake showed an increased association with the prevalence of UL (Q4 vs. Q1: OR_2_ 1.70, 95 % CI 1.02-2.84; *p* trend = 0.049). Higher poultry intake in Q2 and Q4 was associated with an increased prevalence of UL compared to Q1 (Q2 vs. Q1: OR_2_ 1.81, 95% CI 1.05- 3.12; Q4 vs. Q1: OR_2_ 1.85, 95% CI 1.09-3.14), although the dose-response trend was not statistically significant (*p* for trend = 0.07).

**Table 2.**
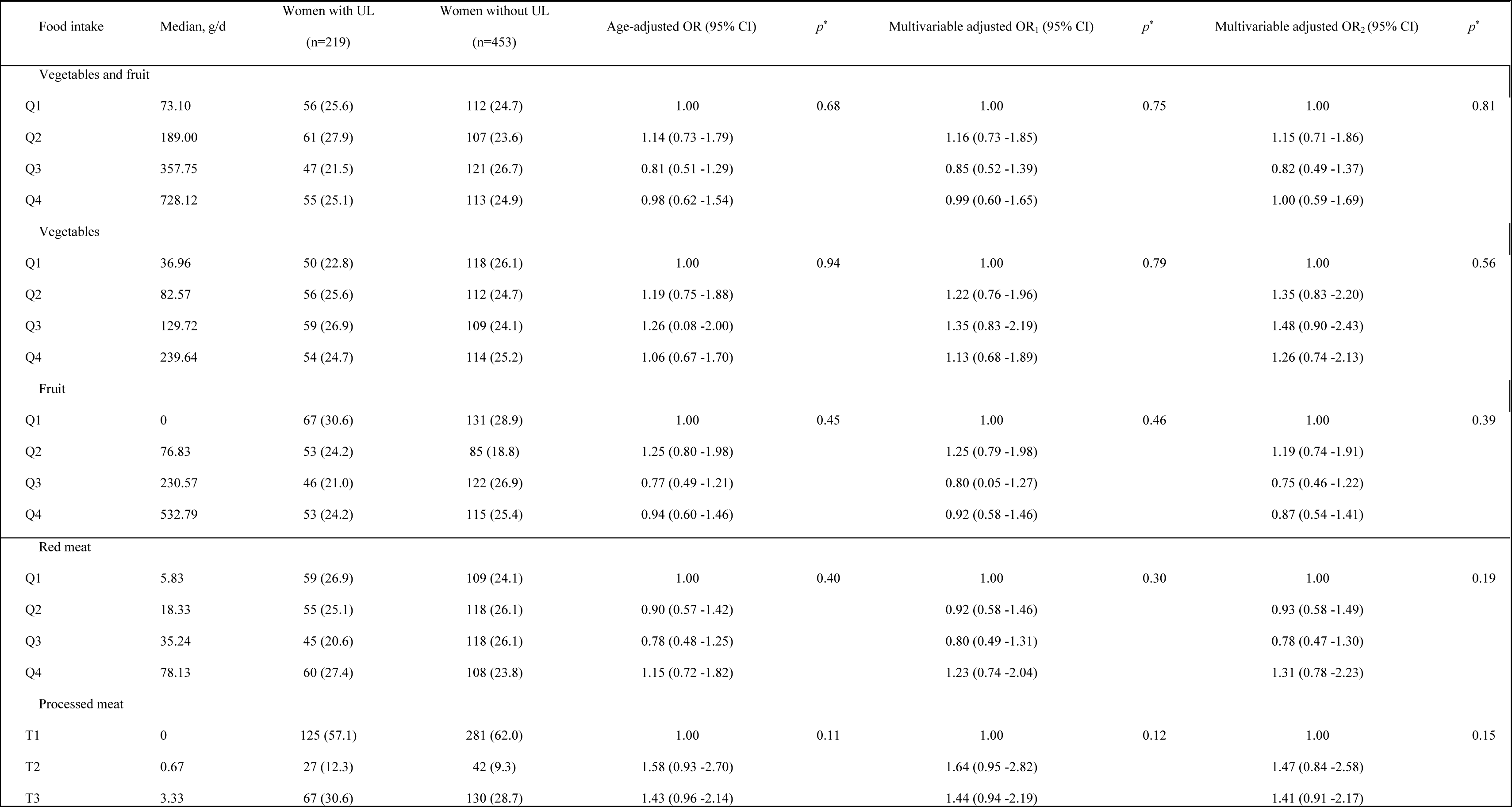
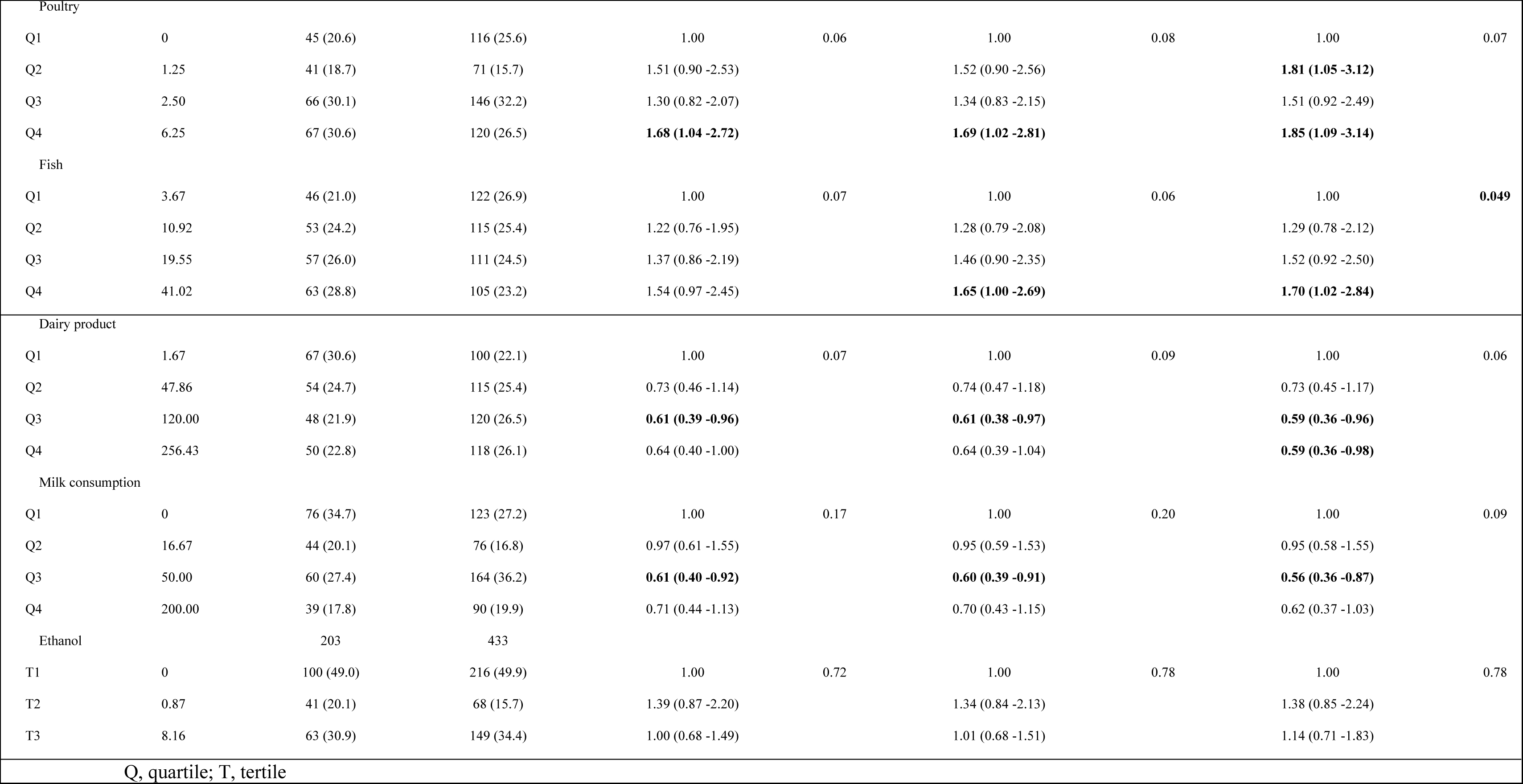
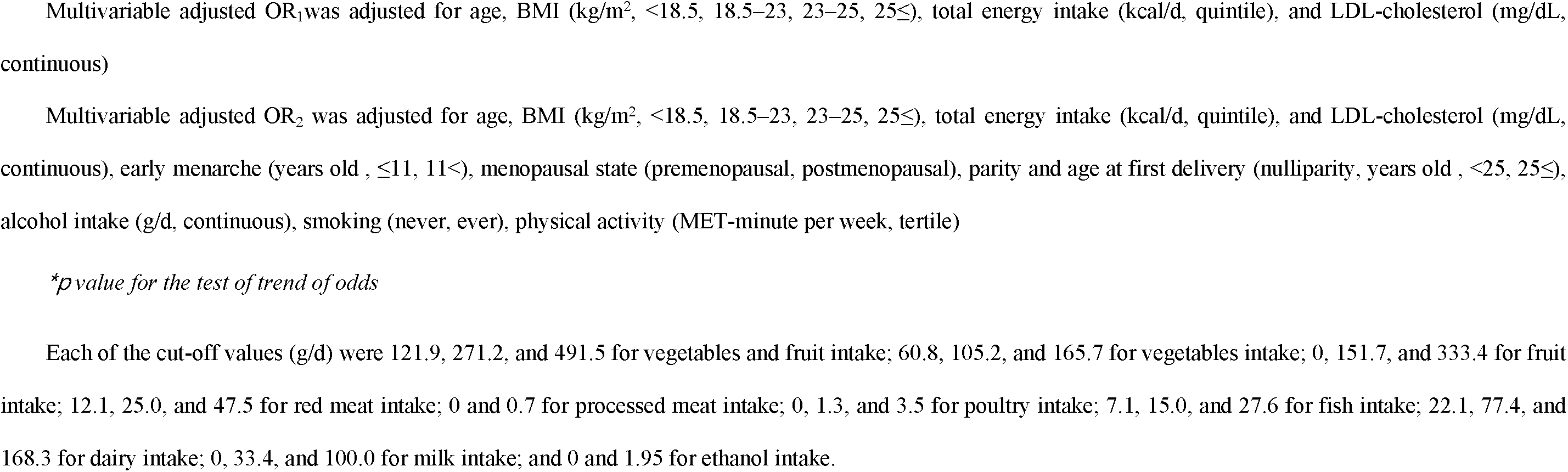
Odds ratios (ORs) and 95% confidence intervals (CIs) of the risk of uterine leiomyoma according to quartiles of intake of each food group in all participants.

On the other hand, higher intake of dairy products in Q3 and Q4 exhibited a significant inverse association with the prevalence of UL compared to Q1 (Q3 vs. Q1: OR_2_ 0.59, 95% CI 0.36-0.96; Q4 vs. Q1: OR_2_ 0.59, 95% CI 0.36-0.98), but the dose-response relationship was not statistically significant (*p* trend = 0.06). The Q3 intake of milk showed a significant inverse association compared to Q1 (Q3 vs. Q1: OR_2_ 0.56, 95% CI 0.36-0.87).

### Association between dietary intake and the prevalence of UL in subgroups according to menopausal status

A subgroup analysis was conducted for pre-and postmenopausal women (Table 3, Figure 1). In premenopausal women, vegetable intake was significantly inversely association with the UL prevalence in a dose-dependent manner (*p* trend = 0.01), although no single quartile intake of vegetables reached statistical significance compared with the lowest quartile (Q4 vs. Q1; OR_2_ 0.47, 95% CI 0.22-1.01). Intake of red meat (Q2 vs. Q1: OR_2_ 0.47, 95% CI 0.24-0.91) and dairy products (Q3 vs. Q1: OR_2_ 0.46, 95% CI 0.23-0.93) showed an inverse association with UL prevalence. The intake of fish was significantly association with increased prevalence of UL (Q3 vs. Q1: OR_2_ 2.83, 95% CI 1.38-5.78). In postmenopausal women, higher fish intake was significantly associated with higher UL prevalence (*p* trend = 0.02), although no statistically significant association was demonstrated in each quartile group. Intake of processed meat (top vs. bottom: OR_2_ 2.33, 95% CI 1.21-4.49) and poultry (Q2 vs. Q1: OR_2_ 2.17, 95% CI 1.04-4.51) showed a significant association with an increased UL prevalence.

**Figure 1.**
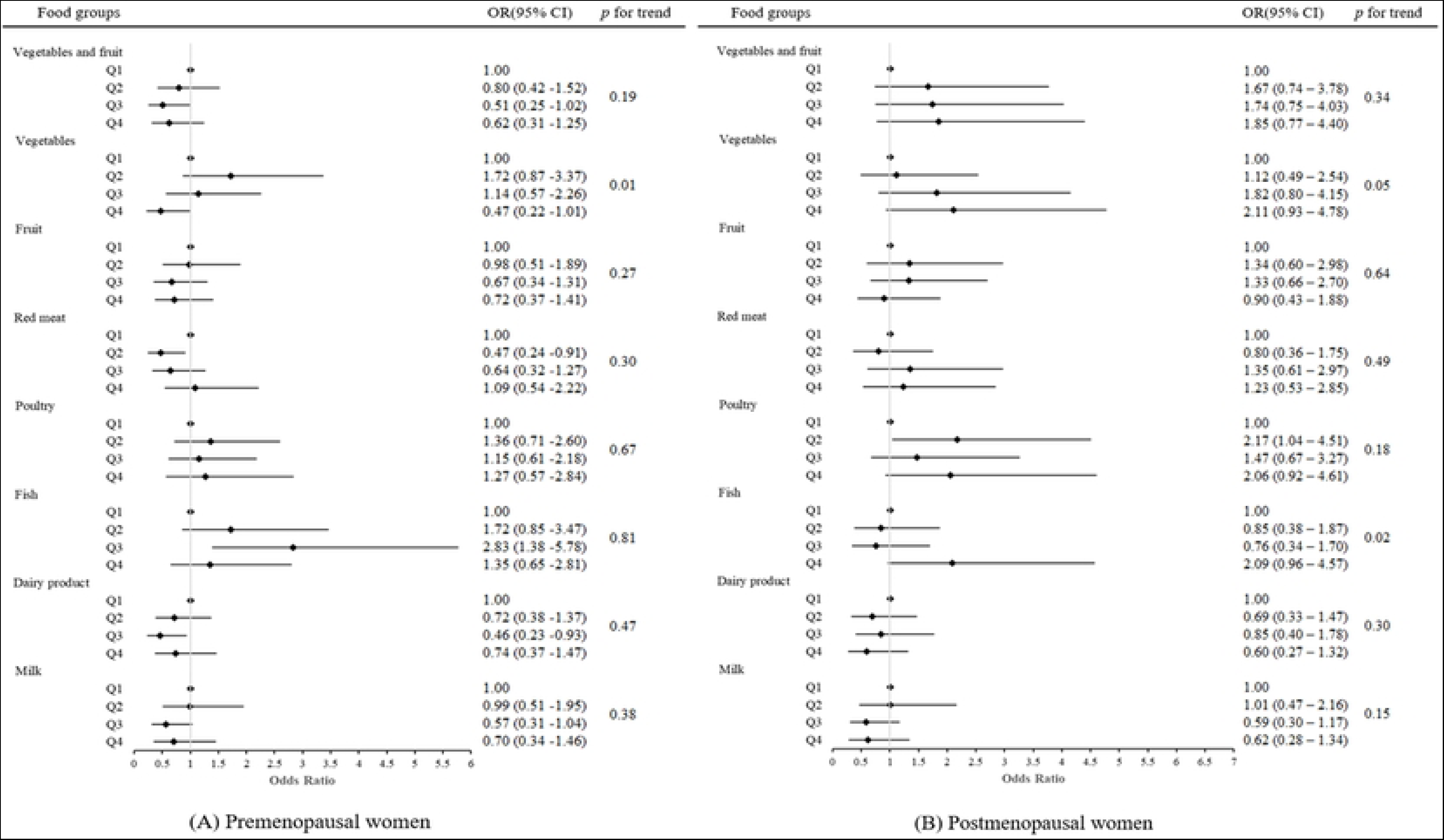
**Association between dietary intake and the prevalence of UL in subgroups according to menopausal status.**

**Table 3.**
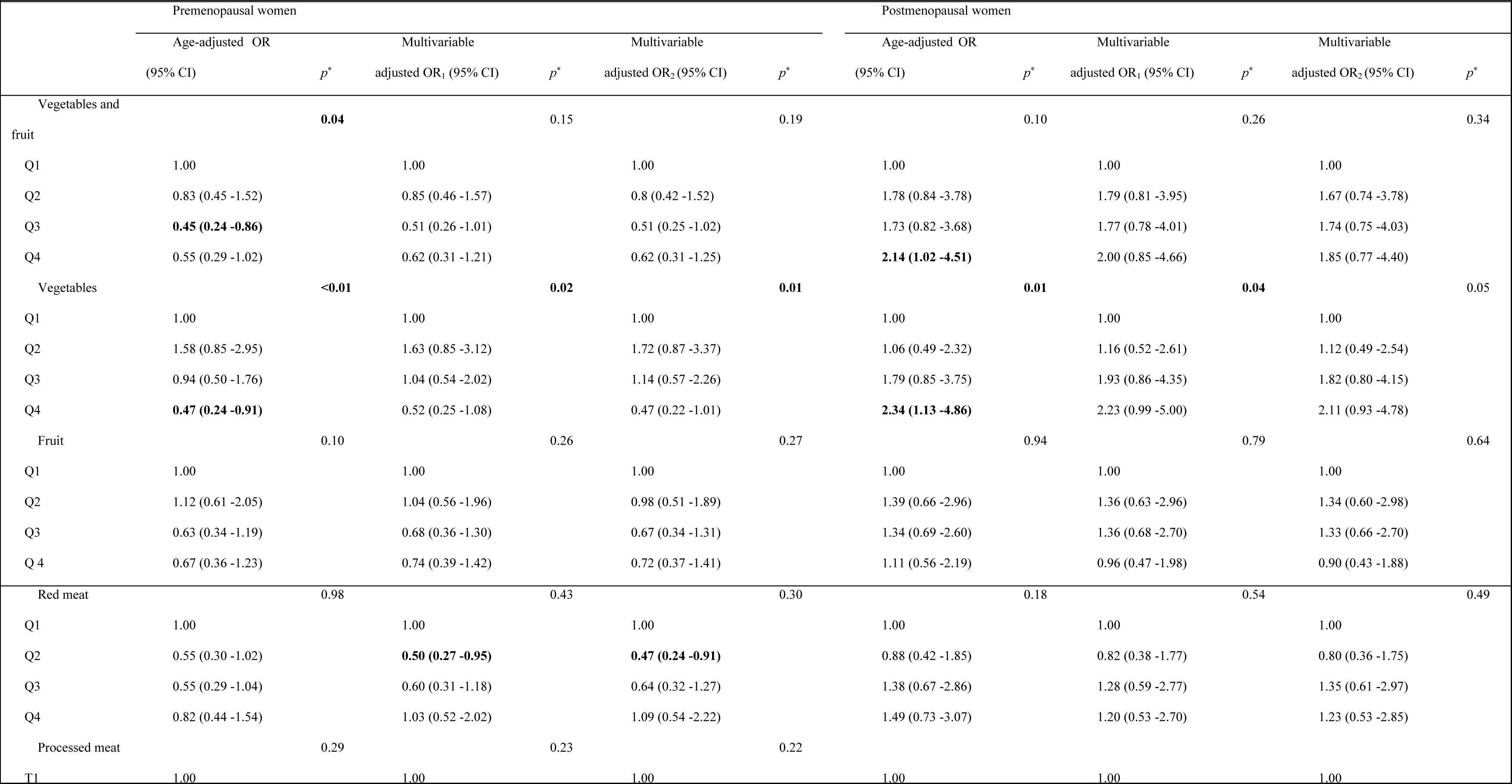
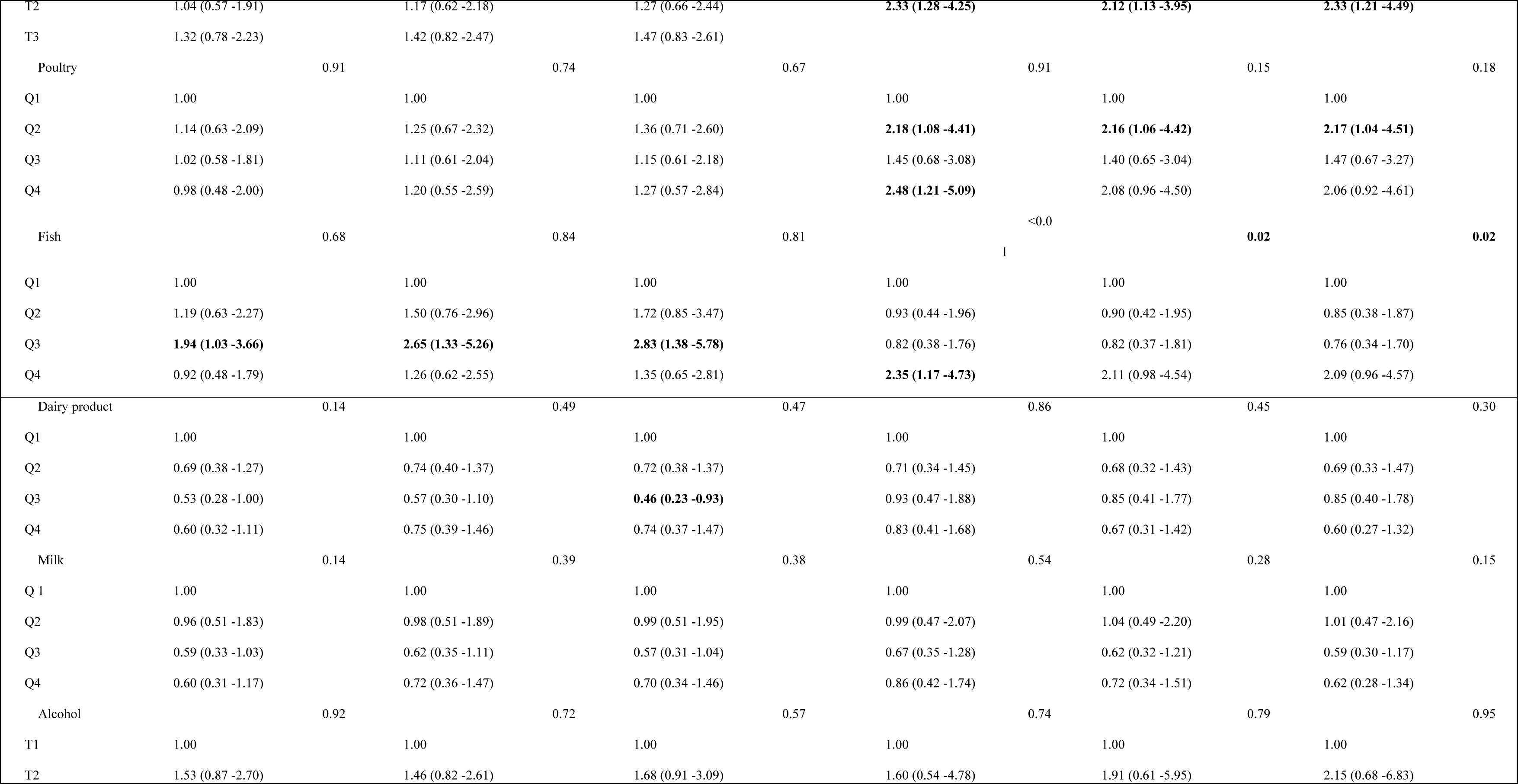
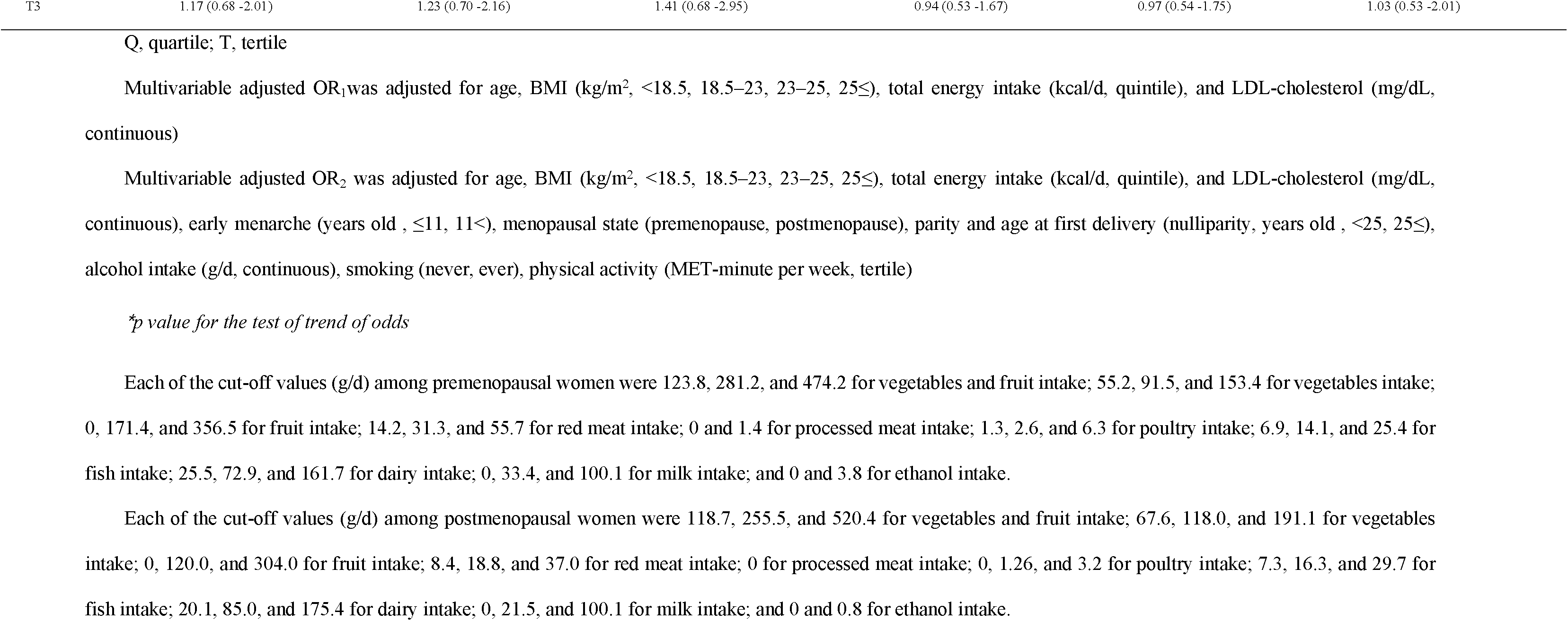
Odds ratios (ORs) and 95% confidence intervals (CIs) of the risk of uterine leiomyoma according to quartiles of intake of each food group in pre-and postmenopausal women.

## Discussion

### Principal findings of our study and results of the other studies

The key findings of our study are as follows:

1. Among all participants, 219 (32.6%) were diagnosed with UL: 132 out of 383 (34.5%) pre-menopausal women and 87 out of 289 (30.1%) postmenopausal women. No statistically significant differences was observed in the prevalence of UL between pre-and postmenopausal women (*p* = 0.23).
2. Elevated fish and poultry consumption were associated with a higher prevalence of UL, with odds ratios (95% confidence intervals) comparing the top vs. bottom quartiles of 1.70 (1.02-2.84; *p* trend = 0.049) for fish intake and 1.85 (1.09 -3.14; *p* trend = 0.07) for poultry intake. Conversely, a higher intake of dairy products displayed an inverse association with UL prevalence (OR 0.59, 95% CI 0.36–0.98; *p* trend = 0.06).
3. Upon analyzing pre-and post-menopausal women separately, a similar pattern emerged with increased prevalence associated with fish intake and decreased prevalence linked to dairy product intake. However, the association between poultry intake and UL prevalence was mainly evident among postmenopausal women. Among premenopausal women, a higher vegetable intake was associated with a lower prevalence of UL (OR 0.47, 95% CI 0.22 -1.01 for top vs. bottom quartiles; *p* trend = 0.01).

Our study observed a 33 % prevalence of UL. Although not statistically significant, premenopausal women exhibited a slightly higher UL prevalence than postmenopausal women (35 % vs. 30 %). Approximately 50% of ULs are asymptomatic and can be accurately diagnosed through ultrasound [1, 24]. We employed rigorous diagnostic criteria for identifying ULs. As a result, ULs were confirmed in about one-third of the total population, a figure comparable to a prior Korean study where ULs were detected in 37.5% of individuals undergoing pelvic ultrasonography as part of health check-ups, regardless of symptom presence [25].

We observed a significant association between fish consumption and UL in premenopausal women (Q3 vs. Q1: OR2 2.83, 95% CI 1.38-5.78) and a dose-dependent association in postmenopausal women (Q4 vs. Q1: OR2 2.09, 95% CI 0.96-4.57; p trend = 0.02). Contrasting findings exist in the literature, with an Italian case-control study reporting an inverse association [26], while Chinese and Japanese studies found no significant differences [27, 28]. A US cohort study demonstrated a 1.2-fold increased prevalence of UL in women who consumed sport fish from the Great Lakes for a decade, suggesting a potential risk elevation due to polychlorinated biphenyls (PCBs) exposure from fish consumption [29]. However, a recent prospective cohort study investigating the relationship between PCBs and UL found no significant correlation [30]. Most previous studies have predominantly analyzed fish consumption in terms of dietary fat [15]. For instance, a prospective study involving 1,171 premenopausal African-American women in the US indicated that intakes of total fat, saturated fat, monounsaturated fat, polyunsaturated fat, and trans-fat were not appreciably associated with UL incidence. Interestingly, the consumption of marine ω-3 polyunsaturated fatty acid, specifically docosahexaenoic acid, was linked to a 49% higher UL incidence (Q4 vs. Q1: HR 1.49, 95% CI 1.04, 2.14, *p* trend = 0.01) [31]. Nevertheless, based solely on the results of this study, it is challenging to estimate the nutritional components of fish contributing to the association between fish consumption and UL. This point applies to all the associations between food groups included in this study and UL, and it has been reiterated as a limitation of this research.

Vegetable intake demonstrated a significant protective association with the prevalence of UL in premenopausal women. The odds ratio of the highest quartile of vegetable intake compared to the bottom quartile was 0.47 (95% CI 0.22–1.01), with a significant dose-dependent relationship (*p* trend = 0.01). These findings align with previous research, such as the Black Women’s Health Study, indicating reduced risk of UL development with higher fruit and vegetable consumption (four or more servings of fruits or vegetables daily, IRR = 0.90, 95% CI 0.82–0.98) [32]. In a case-control study involving 273 women, of whom 94% were of Han Chinese ethnicity, a negative correlation was found between vegetable and fruit intake and UL (OR 0.5, 95% CI 0.3–0.9) in premenopausal women [27]. Furthermore, other investigations have indicated that women with UL consume green vegetables and fruits less frequently than women without UL [26, 33]. These protective associations are attributed to mechanisms such as decreased bioavailable estrogen and growth factors [34, 35], or elevated levels of phytochemicals with anti-inflammatory properties [36, 37]. However, this study did not observe a protective effect of fruit consumption. The odds ratios for combined vegetable and fruit intake, as well as fruit intake alone, were 0.62 (95% CI 0.31–1.25) and 0.72 (95% CI 0.37–1.41) for the highest quartile over the lowest quartile, respectively. This finding aligns with a case-control study involving 843 Italian women, which indicated that vegetables were more protective than fruits against UL prevalence (OR 0.5, 95% CI 0.4–0.6 for green vegetables, OR 0.8, 95% CI 0.6–1.0 for fruit consumption) [26].

We observed a protective association between dairy consumption and UL prevalence (Q4 vs. Q1: OR_2_ 0.59, 95% CI 0.36-0.98 for all participants; Q3 vs. Q1: OR_2_ 0.46, 95% CI 0.23-0.93 for premenopausal women). These findings align with a previous study that reported a protective effect of frequent consumption of milk and low-fat dairy products, as well as a modest protective effect for yogurt consumption, against the occurrence of UL. However, no significant associations were found for butter, cheese, and ice cream among African American women [38]. On the contrary, an Italian study presented contrasting results, finding no association between milk and cheese intake and the risk of UL [26]. Additionally, a Chinese prospective cohort study demonstrated an increased risk when analyzing combined milk and soymilk consumption [39]. Dairy products are complex compounds with composition variations influenced by regional disparities in livestock production environments. Furthermore, reports suggest that dairy products might contain estrogenic compounds that could be absorbed and affect the menstrual cycle [40, 41]. As a result, diverse research findings have emerged concerning the link between dairy consumption and the risk of UL. In a substantial prospective cohort study spanning 18 years, no distinct associations emerged between overall dairy consumption and the risk of UL. Nevertheless, the study did establish that yogurt intake and dietary calcium were associated with a reduced risk of UL development [42].

Regarding the connection between meat consumption and UL, we observed a protective association with certain levels of red meat intake in premenopausal women (Q2 vs. Q1: OR_2_ 0.47, 95% CI 0.24-0.91). On the other hand, we identified an increased association between processed meat (higher vs. lower: OR_2_ 2.33, 95% CI 1.21-4.49) and poultry (Q2 vs. Q1: OR_2_ 2.17, 95% CI 1.04-4.51) consumption and UL prevalence in postmenopausal women. This aligns with findings from an Italian case-control study, which demonstrated that significant consumption of meats such as beef or ham was associated with an elevated risk of UL [26]. However, this risk was found to be insignificant in the Chinese population [27]. To classify meats, we categorized them into red meat, processed meat, and poultry. Processed meat and poultry intake exhibited an association with increased UL prevalence exclusively in postmenopausal women, whereas red meat intake indicated a lower UL prevalence solely in premenopausal women. Interpreting these associations is limited by the relatively low absolute amount of meat intake within this population. Nonetheless, it appears that processed meat, rather than red meat, may contain specific metabolites that could stimulate proliferative activities in UL cells [43, 44].

A notable finding in our study was the variation in the association between dietary patterns and UL prevalence based on menopausal status. Numerous studies have reported differing dietary impacts on hormone-related conditions contingent on menopausal status. For instance, an investigation into the impact of a diabetes risk reduction diet on endometrial cancer revealed inverse associations exclusively among postmenopausal women, without such effects seen in premenopausal women [45]. Similarly, a study exploring the link between urinary isoflavone and urinary estrogen levels after isoflavone intake identified a positive correlation solely in postmenopausal women. Cumulative evidence from epidemiological and metabolomics research suggests that the postmenopausal state can influence a specific set of metabolites in response to a particular diet, distinct from the premenopausal state [46, 47]. The potential anti-proliferative effect of a diet might be more profound in the high estrogenic environment of premenopausal women. Conversely, the impact of certain dietary components, such as estrogenic compounds in fatty fish or processed meat, could be more significant in the hypoestrogenic context of postmenopausal women. Consequently, considering the influence of menopausal status is imperative when investigating the relationship between dietary intake and health outcomes.

### Strengths and limitations

To the best of our knowledge, this is the first Korean study to investigate the association between dietary factors and UL, employing a validated Food Frequency Questionnaire (FFQ) known for its commendable reproducibility and validity [20]. All UL cases and non-cases underwent diagnosis through pelvic ultrasound examination, considered the most sensitive and clinically effective diagnostic tool for UL. Notably, the assessment of dietary intake data coincided with clinical factors, with the analysis accounting for menopausal status and other relevant confounding factors. However, several limitations must be acknowledged. Firstly, inevitable information discrepancies may arise from data gathered through self-reported food intake questionnaires. Secondly, the cross-sectional nature of the study curtails the ability to infer causal relationships, raising the possibility that those already diagnosed with UL may exhibit specific dietary patterns. Furthermore, the study may be subject to potential bias toward individuals of medium to high socioeconomic status who willingly invested USD $500–1300 for private health assessments, influencing the findings due to their increased health awareness and motivation towards adopting healthier lifestyles. Thirdly, the single-center design warrants caution in generalizing these findings to the entirety of Korean women. Fourthly, the study did not delve into the impact of nutrients derived from foods on dietary intake analyses. Absent were data on the concentration of associated metabolites, blood markers of inflammation, and reproductive or growth hormones, presumed to mediate these associations. Lastly, this study excluded women who had undergone hysterectomy due to UL-related symptoms, inadvertently excluding severe or symptomatic cases.

## Conclusions

In our study involving Korean women who underwent pelvic ultrasonography, we found that high consumption of fish and poultry, coupled with low intake of dairy products, correlated with an elevated prevalence of UL. Furthermore, vegetable intake exhibited an inverse relationship with UL prevalence, particularly among premenopausal women. These results suggest that dietary interventions offer promise as a potential preventive strategy for UL, with a specific focus on premenopausal women who are disproportionately affected by this prevalent and consequential gynecological condition.

## Data Availability

All relevant data are within the manuscript and its Supporting Information files.

## Supporting data

**Supplementary Table 1. Median values of the tertiles or quartiles of each dietary group in all, pre-, and postmenopausal women.**

## References

1. Baird DD, Dunson DB, Hill MC, Cousins D, Schectman JM. High cumulative incidence of uterine leiomyoma in black and white women: ultrasound evidence. Am J Obstet Gynecol. 2003;188(1):100–7. doi: 10.1067/mob.2003.99. PubMed PMID: 12548202.

2. Stewart EA, Cookson CL, Gandolfo RA, Schulze-Rath R. Epidemiology of uterine fibroids: a systematic review. Bjog. 2017;124(10):1501–12. Epub 20170513. doi: 10.1111/1471-0528.14640. PubMed PMID: 28296146.

3. Wise LA, Laughlin-Tommaso SK. Epidemiology of Uterine Fibroids: From Menarche to Menopause. Clin Obstet Gynecol. 2016;59(1):2–24. doi: 10.1097/grf.0000000000000164. PubMed PMID: 26744813; PubMed Central PMCID: PMCPMC4733579.

4. Bulun SE. Uterine fibroids. N Engl J Med. 2013;369(14):1344-55. doi: 10.1056/NEJMra1209993. PubMed PMID: 24088094.

5. Bulun SE, Moravek MB, Yin P, Ono M, Coon JSt, Dyson MT, et al. Uterine Leiomyoma Stem Cells: Linking Progesterone to Growth. Semin Reprod Med. 2015;33(5):357–65. Epub 20150806. doi: 10.1055/s-0035-1558451. PubMed PMID: 26251118.

6. Hunter DS, Hodges LC, Eagon PK, Vonier PM, Fuchs-Young R, Bergerson JS, et al. Influence of exogenous estrogen receptor ligands on uterine leiomyoma: evidence from an in vitro/in vivo animal model for uterine fibroids. Environ Health Perspect. 2000;108 Suppl 5:829–34. doi: 10.1289/ehp.00108s5829. PubMed PMID: 11035990.

7. Marsh EE, Bulun SE. Steroid hormones and leiomyomas. Obstet Gynecol Clin North Am. 2006;33(1):59–67. doi: 10.1016/j.ogc.2005.12.001. PubMed PMID: 16504806.

8. Afrin S, AlAshqar A, El Sabeh M, Miyashita-Ishiwata M, Reschke L, Brennan JT, et al. Diet and Nutrition in Gynecological Disorders: A Focus on Clinical Studies. Nutrients. 2021;13(6). Epub 20210521. doi: 10.3390/nu13061747. PubMed PMID: 34063835; PubMed Central PMCID: PMCPMC8224039.

9. Pavone D, Clemenza S, Sorbi F, Fambrini M, Petraglia F. Epidemiology and Risk Factors of Uterine Fibroids. Best Pract Res Clin Obstet Gynaecol. 2018;46:3–11. Epub 20171001. doi: 10.1016/j.bpobgyn.2017.09.004. PubMed PMID: 29054502.

10. Bradbury KE, Appleby PN, Key TJ. Fruit, vegetable, and fiber intake in relation to cancer risk: findings from the European Prospective Investigation into Cancer and Nutrition (EPIC). Am J Clin Nutr. 2014;100 Suppl 1:394s–8s. Epub 20140611. doi: 10.3945/ajcn.113.071357. PubMed PMID: 24920034.

11. Friberg E, Wallin A, Wolk A. Sucrose, high-sugar foods, and risk of endometrial cancer--a population-based cohort study. Cancer Epidemiol Biomarkers Prev. 2011;20(9):1831–7. Epub 20110715. doi: 10.1158/1055-9965.Epi-11-0402. PubMed PMID: 21765006.

12. Masala G, Assedi M, Bendinelli B, Ermini I, Sieri S, Grioni S, et al. Fruit and vegetables consumption and breast cancer risk: the EPIC Italy study. Breast Cancer Res Treat. 2012;132(3):1127–36. Epub 20120104. doi: 10.1007/s10549-011-1939-7. PubMed PMID: 22215387.

13. Rieck G, Fiander A. The effect of lifestyle factors on gynaecological cancer. Best Pract Res Clin Obstet Gynaecol. 2006;20(2):227–51. doi: 10.1016/j.bpobgyn.2005.10.010. PubMed PMID: 16543119.

14. Islam MS, Segars JH, Castellucci M, Ciarmela P. Dietary phytochemicals for possible preventive and therapeutic option of uterine fibroids: Signaling pathways as target. Pharmacol Rep. 2017;69(1):57–70. Epub 20161020. doi: 10.1016/j.pharep.2016.10.013. PubMed PMID: 27898339.

15. Ciebiera M, Esfandyari S, Siblini H, Prince L, Elkafas H, Wojtyła C, et al. Nutrition in Gynecological Diseases: Current Perspectives. Nutrients. 2021;13(4). Epub 20210402. doi: 10.3390/nu13041178. PubMed PMID: 33918317; PubMed Central PMCID: PMCPMC8065992.

16. Parazzini F, Di Martino M, Candiani M, Viganò P. Dietary components and uterine leiomyomas: a review of published data. Nutr Cancer. 2015;67(4):569–79. Epub 20150331. doi: 10.1080/01635581.2015.1015746. PubMed PMID: 25826470.

17. Tinelli A, Vinciguerra M, Malvasi A, Andjić M, Babović I, Sparić R. Uterine Fibroids and Diet. Int J Environ Res Public Health. 2021;18(3). Epub 20210125. doi: 10.3390/ijerph18031066. PubMed PMID: 33504114; PubMed Central PMCID: PMCPMC7908561.

18. Yang SY, Kim YS, Lee JE, Seol J, Song JH, Chung GE, et al. Dietary protein and fat intake in relation to risk of colorectal adenoma in Korean. Medicine (Baltimore). 2016;95(49):e5453. doi: 10.1097/md.0000000000005453. PubMed PMID: 27930524; PubMed Central PMCID: PMCPMC5265996.

19. Harlow SD, Gass M, Hall JE, Lobo R, Maki P, Rebar RW, et al. Executive summary of the Stages of Reproductive Aging Workshop + 10: addressing the unfinished agenda of staging reproductive aging. J Clin Endocrinol Metab. 2012;97(4):1159–68. Epub 20120216. doi: 10.1210/jc.2011-3362. PubMed PMID: 22344196; PubMed Central PMCID: PMCPMC3319184.

20. Ahn Y, Kwon E, Shim JE, Park MK, Joo Y, Kimm K, et al. Validation and reproducibility of food frequency questionnaire for Korean genome epidemiologic study. Eur J Clin Nutr. 2007;61(12):1435–41. Epub 20070207. doi: 10.1038/sj.ejcn.1602657. PubMed PMID: 17299477.

21. Alberti KG, Eckel RH, Grundy SM, Zimmet PZ, Cleeman JI, Donato KA, et al. Harmonizing the metabolic syndrome: a joint interim statement of the International Diabetes Federation Task Force on Epidemiology and Prevention; National Heart, Lung, and Blood Institute; American Heart Association; World Heart Federation; International Atherosclerosis Society; and International Association for the Study of Obesity. Circulation. 2009;120(16):1640–5. Epub 20091005. doi: 10.1161/circulationaha.109.192644. PubMed PMID: 19805654.

22. Lee SY, Park HS, Kim DJ, Han JH, Kim SM, Cho GJ, et al. Appropriate waist circumference cutoff points for central obesity in Korean adults. Diabetes Res Clin Pract. 2007;75(1):72–80. Epub 20060602. doi: 10.1016/j.diabres.2006.04.013. PubMed PMID: 16735075.

23. Chun MY. Validity and reliability of korean version of international physical activity questionnaire short form in the elderly. Korean J Fam Med. 2012;33(3):144–51. Epub 20120524. doi: 10.4082/kjfm.2012.33.3.144. PubMed PMID: 22787536; PubMed Central PMCID: PMCPMC3391639.

24. Foth D, Röhl FW, Friedrich C, Tylkoski H, Rabe T, Römer T, et al. Symptoms of uterine myomas: data of an epidemiological study in Germany. Arch Gynecol Obstet. 2017;295(2):415–26. Epub 20161121. doi: 10.1007/s00404-016-4239-y. PubMed PMID: 27873052.

25. Kim MH, Park YR, Lim DJ, Yoon KH, Kang MI, Cha BY, et al. The relationship between thyroid nodules and uterine fibroids. Endocr J. 2010;57(7):615–21. Epub 20100513. doi: 10.1507/endocrj.k10e-024. PubMed PMID: 20467159.

26. Chiaffarino F, Parazzini F, La Vecchia C, Chatenoud L, Di Cintio E, Marsico S. Diet and uterine myomas. Obstet Gynecol. 1999;94(3):395-8. doi: 10.1016/s0029-7844(99)00305-1. PubMed PMID: 10472866.

27. He Y, Zeng Q, Dong S, Qin L, Li G, Wang P. Associations between uterine fibroids and lifestyles including diet, physical activity and stress: a case-control study in China. Asia Pac J Clin Nutr. 2013;22(1):109–17. doi: 10.6133/apjcn.2013.22.1.07. PubMed PMID: 23353618.

28. Nagata C, Nakamura K, Oba S, Hayashi M, Takeda N, Yasuda K. Association of intakes of fat, dietary fibre, soya isoflavones and alcohol with uterine fibroids in Japanese women. Br J Nutr. 2009;101(10):1427–31. doi: 10.1017/s0007114508083566. PubMed PMID: 19459228.

29. Lambertino A, Turyk M, Anderson H, Freels S, Persky V. Uterine leiomyomata in a cohort of Great Lakes sport fish consumers. Environ Res. 2011;111(4):565–72. Epub 20110209. doi: 10.1016/j.envres.2011.01.006. PubMed PMID: 21310402; PubMed Central PMCID: PMCPMC3111144.

30. Wesselink AK, Claus Henn B, Fruh V, Orta OR, Weuve J, Hauser R, et al. A Prospective Ultrasound Study of Plasma Polychlorinated Biphenyl Concentrations and Incidence of Uterine Leiomyomata. Epidemiology. 2021;32(2):259–67. doi: 10.1097/ede.0000000000001320. PubMed PMID: 33427764; PubMed Central PMCID: PMCPMC8862183.

31. Brasky TM, Bethea TN, Wesselink AK, Wegienka GR, Baird DD, Wise LA. Dietary Fat Intake and Risk of Uterine Leiomyomata: A Prospective Ultrasound Study. Am J Epidemiol. 2020;189(12):1538–46. doi: 10.1093/aje/kwaa097. PubMed PMID: 32556077; PubMed Central PMCID: PMCPMC7857646.

32. Wise LA, Radin RG, Palmer JR, Kumanyika SK, Boggs DA, Rosenberg L. Intake of fruit, vegetables, and carotenoids in relation to risk of uterine leiomyomata. Am J Clin Nutr. 2011;94(6):1620–31. Epub 20111109. doi: 10.3945/ajcn.111.016600. PubMed PMID: 22071705; PubMed Central PMCID: PMCPMC3252555.

33. Shen Y, Wu Y, Lu Q, Ren M. Vegetarian diet and reduced uterine fibroids risk: A case-control study in Nanjing, China. J Obstet Gynaecol Res. 2016;42(1):87–94. Epub 20151012. doi: 10.1111/jog.12834. PubMed PMID: 26458740.

34. Allen NE, Appleby PN, Davey GK, Kaaks R, Rinaldi S, Key TJ. The associations of diet with serum insulin-like growth factor I and its main binding proteins in 292 women meat-eaters, vegetarians, and vegans. Cancer Epidemiol Biomarkers Prev. 2002;11(11):1441–8. PubMed PMID: 12433724.

35. Barnard ND, Scialli AR, Hurlock D, Bertron P. Diet and sex-hormone binding globulin, dysmenorrhea, and premenstrual symptoms. Obstet Gynecol. 2000;95(2):245–50. doi: 10.1016/s0029-7844(99)00525-6. PubMed PMID: 10674588.

36. Armstrong BK, Brown JB, Clarke HT, Crooke DK, Hähnel R, Masarei JR, et al. Diet and reproductive hormones: a study of vegetarian and nonvegetarian postmenopausal women. J Natl Cancer Inst. 1981;67(4):761–7. PubMed PMID: 6944545.

37. Islam MS, Akhtar MM, Ciavattini A, Giannubilo SR, Protic O, Janjusevic M, et al. Use of dietary phytochemicals to target inflammation, fibrosis, proliferation, and angiogenesis in uterine tissues: promising options for prevention and treatment of uterine fibroids? Mol Nutr Food Res. 2014;58(8):1667–84. Epub 20140630. doi: 10.1002/mnfr.201400134. PubMed PMID: 24976593; PubMed Central PMCID: PMCPMC4152895.

38. Wise LA, Radin RG, Palmer JR, Kumanyika SK, Rosenberg L. A prospective study of dairy intake and risk of uterine leiomyomata. Am J Epidemiol. 2010;171(2):221–32. Epub 20091202. doi: 10.1093/aje/kwp355. PubMed PMID: 19955473; PubMed Central PMCID: PMCPMC2800240.

39. Gao M, Wang H. Frequent milk and soybean consumption are high risks for uterine leiomyoma: A prospective cohort study. Medicine (Baltimore). 2018;97(41):e12009. doi: 10.1097/md.0000000000012009. PubMed PMID: 30313022; PubMed Central PMCID: PMCPMC6203589.

40. Kim K, Wactawski-Wende J, Michels KA, Plowden TC, Chaljub EN, Sjaarda LA, et al. Dairy Food Intake Is Associated with Reproductive Hormones and Sporadic Anovulation among Healthy Premenopausal Women. J Nutr. 2017;147(2):218–26. Epub 20161123. doi: 10.3945/jn.116.241521. PubMed PMID: 27881593; PubMed Central PMCID: PMCPMC5265695.

41. Maruyama K, Oshima T, Ohyama K. Exposure to exogenous estrogen through intake of commercial milk produced from pregnant cows. Pediatr Int. 2010;52(1):33–8. Epub 20090522. doi: 10.1111/j.1442-200X.2009.02890.x. PubMed PMID: 19496976.

42. Orta OR, Terry KL, Missmer SA, Harris HR. Dairy and related nutrient intake and risk of uterine leiomyoma: a prospective cohort study. Hum Reprod. 2020;35(2):453–63. doi: 10.1093/humrep/dez278. PubMed PMID: 32086510; PubMed Central PMCID: PMCPMC8489562.

43. Chajès V, Thiébaut AC, Rotival M, Gauthier E, Maillard V, Boutron-Ruault MC, et al. Association between serum trans-monounsaturated fatty acids and breast cancer risk in the E3N-EPIC Study. Am J Epidemiol. 2008;167(11):1312–20. Epub 20080404. doi: 10.1093/aje/kwn069. PubMed PMID: 18390841; PubMed Central PMCID: PMCPMC2679982.

44. Turesky RJ. Mechanistic Evidence for Red Meat and Processed Meat Intake and Cancer Risk: A Follow-up on the International Agency for Research on Cancer Evaluation of 2015. Chimia (Aarau). 2018;72(10):718–24. doi: 10.2533/chimia.2018.718. PubMed PMID: 30376922; PubMed Central PMCID: PMCPMC6294997.

45. Esposito G, Bravi F, Serraino D, Parazzini F, Crispo A, Augustin LSA, et al. Diabetes Risk Reduction Diet and Endometrial Cancer Risk. Nutrients. 2021;13(8). Epub 20210730. doi: 10.3390/nu13082630. PubMed PMID: 34444790; PubMed Central PMCID: PMCPMC8399314.

46. Stark KD, Park EJ, Holub BJ. Fatty acid composition of serum phospholipid of premenopausal women and postmenopausal women receiving and not receiving hormone replacement therapy. Menopause. 2003;10(5):448–55. doi: 10.1097/01.Gme.0000059861.93639.1a. PubMed PMID: 14501607.

47. Verri Hernandes V, Dordevic N, Hantikainen EM, Sigurdsson BB, Smárason SV, Garcia-Larsen V, et al. Age, Sex, Body Mass Index, Diet and Menopause Related Metabolites in a Large Homogeneous Alpine Cohort. Metabolites. 2022;12(3). Epub 20220224. doi: 10.3390/metabo12030205. PubMed PMID: 35323648; PubMed Central PMCID: PMCPMC8955763.

